# PERFORMANCE EVALUATION OF A SARS-COV-2 RAPID ANTIGENTEST: TEST PERFORMANCE IN THE COMMUNITY IN THE NETHERLANDS

**DOI:** 10.1101/2020.10.19.20215202

**Authors:** N. Van der Moeren, V.F. Zwart, E.B. Lodder, W. Van den Bijllaardt, H.R.J.M. Van Esch, J.J.J.M. Stohr, J. Pot, I. Welschen, P.M.F. Van Mechelen, S.D. Pas, J.A.J.W. Kluytmans

## Abstract

**Objectives:** This study was primarily conducted to evaluate clinical sensitivity and specificity of the SARS-CoV-2 rapid antigen test ‘BD Veritor System for Rapid Detection of SARS-CoV-2’ (VRD) compared to real time reverse transcriptase polymerase chain reaction (qRT-PCR). Furthermore, the VRD sensitivity for different Ct-value groups (Ct <20; Ct 20-25, Ct 25-30 and Ct ≥30) and different intervals since symptom onset (< 7 days; ≥ 7 days) were examined.

**Design:** Prospective performance evaluation study.

**Setting:** Municipal Health Service (GGD) COVID-19 test centres in West-Brabant, the Netherlands

**Participants:** In order to evaluate clinical specificity, 352 symptomatic adults (≥18 years) who presented at a participating GGD test centre for a COVID-19 test between September 28 and October 7 2020 were included. In order to evaluate clinical sensitivity, 123 symptomatic adults (≥18 years) who were tested positive with qRT-PCR in a participating GGD test centre between September 26 and October 6 were included.

**Results:** An overall clinical specificity of 100% (95%CI: 98.9%-100%) and sensitivity of 80.7% (95% CI: 73,2%-86,9%) was found for the VRD compared to qRT-PCR. Sensitivity was the highest for low Ct-value categories and for specimen obtained within the first days after disease onset. For specimen obtained within 7 days after onset of symptoms, the overall sensitivity was 91.0% (95% CI: 82,4%-96,3%) and 98,6% (95% CI: 92,3%-100%) for samples with qRT-PCR Ct-value beneath 30.

**Conclusion:** The VRD is a promising diagnostic test for COVID-19 community screening for symptomatic individuals within 7 days after symptom onset in function of disease control. The clinical sensitivity was highest when viral load was high, which correlated with the duration of symptoms. Further research on practical applicability and the optimal position of the test within the current testing landscape is needed.

## Background

Accurate and sustainable test strategies are essential for the control of COVID-19. (1) The current test used to establish COVID-19 infection in The Netherlands is real-time reverse transcriptase polymerase chain reaction (qRT-PCR). This test is highly sensitive and specific and therefore well suited for the diagnosis of clinically ill patients. However, application of the test for large-scale community screening for disease control raises substantial challenges. qRT-PCR can only be performed in specialised laboratories, has a relatively long turnaround time (TAT) and depends on the availability of scarce extraction and PCR reagents and disposables. The massive qRT-PCR demand created by community screening greatly pressurizes microbiological laboratories and puts routine clinical diagnostic care at risk. Furthermore, logistic and administrative challenges lead to substantial delays in testing and reporting of the results. Rapid testing is however key in the control of SARS-CoV-2 community spread. (2) COVID-19 community screening requires a low-cost diagnostic test with a short TAT which can be performed close to the community. Lateral flow assay (LFA) SARS-CoV-2 antigen tests can be performed at point of care, give results within 15-30 minutes and are relatively inexpensive to produce. (3,4) Numerous SARS-CoV-2 LFA are available, but qualitative data on their performance is scarce. The rare available studies are often based on remnant laboratory samples and contain little information on clinical setting or disease stage. The current literature is insufficient to determine whether SARS-CoV-2 rapid antigen test can be useful in clinical practice and prospective evaluation of the antigen tests in clinically relevant settings is needed. (5)

Hence, this clinical performance study of the ‘BD Veritor System for Rapid Detection of SARS-CoV-2’ (VRD), a chromatographic immunoassay for the qualitative detection of nucleocapsid antigens in respiratory specimen, at two COVID-19 test centres of the Dutch Municipal Health Service (GGD).

## Methods

### Objectives

The primary study objective of the study was to determine the clinical specificity and sensitivity of the BD Veritor System for Rapid Detection of SARS-CoV-2 (VRD) compared to qRT-PCR. Secondary objectives were to determine the clinical sensitivity for different Ct-value groups (Ct <20, Ct 20-25, Ct 25-30 and Ct ≥30) and different intervals since symptom onset (< 7 days, ≥ 7 days). Also we aimed to evaluate the concordance between visual interpretation of VRD test results and analysis using the reading device provided by the manufacturer, the BD Veritor Analyser (VA).

### Setting

COVID-19 testing of non-hospitalized symptomatic patients in the Netherlands is coordinated by the Municipal Health Service (GGD). A person with COVID-19 like symptoms has to make an appointment at a regional GGD test centre. A single swab is used to collect specimen from throat and nose and is sent to the microbiological laboratory for qRT-PCR. Individuals with a positive qRT-PCR result are informed by a GGD employee and approached with a questionnaire for the purpose of source- and contact-tracing (BCO).

The study was conducted from September 26^th^ to October 7^th^ 2020 in the region West-Brabant, the Netherlands. The local GGD had three operational test centres during the study, conducting 1200 SARS-CoV-2 qRT-PCRs daily. Samples from two centres (Breda and Roosendaal) were considered for this study.

In the third week of September 2020 5-6% of individuals presenting at a West Brabant GGD test centre had a positive qRT-PCR (data on file).

### BD Veritor System for Rapid Detection of SARS-CoV-2

The ‘BD Veritor System for Rapid Detection of SARS-CoV-2’ (VRD) is a chromatographic lateral flow immunoassay for the qualitative detection of nucleocapsid antigens in respiratory specimen. The manufacturer reports a test specificity of 100% and a sensitivity of 84% compared to qRT-PCR as a gold standard during the first 5 days after disease onset. The test was validated by the manufacturer for use on superficial nasal specimen. Interpretation is prescribed after 15 minutes with a reading device provided by the manufacturer, the BD Veritor Analyser (VA). Nevertheless, a test and control line can be seen by eye. (3)

### Real-time reverse transcriptase PCR

Two CE-IVD labelled qPCR platforms were used: the Cobas 6800 (Roche) and the m2000 (Abbott). Both platforms have two targets: the E-gene and RDRP-gene or N-gen respectively.

### Patient recruitment

In a first part of the study with the primary objective to evaluate clinical test specificity, all adults (≥18 years) presenting at the GGD test centre Breda for a COVID-19 test between September 28 and 30, 2020 were invited to participate. Individuals who were able and willing to give verbal informed consent were included.

In a second part of the study with the primary objective to determine clinical test sensitivity, adults (≥18 years) who had been tested at a GGD test centre between September 26 and October 6 with a positive qRT-PCR were approached by a GGD employee and invited to participate. Individuals who gave informed consent by telephone were visited at home and asked for verbal informed consent a second time.

The study protocol was submitted at the medical ethical board ‘Medical research Ethics Committees United’ (MEC-U) and was granted an exemption of the Dutch medical scientific research act (WMO).

### Study procedure

In the first part of the study, one swab was used to obtain a specimen from the throat and nasal cavity up to the nasal bridge for routine qRT-PCR in accordance with the Dutch national COVID-19 test protocol. In addition to and directly following this first swab, the same GGD employee obtained an additional swab to acquire a specimen from the throat and the superficial nasal cavities (bilateral, 2.5 cm proximal from the nostril) for VRD. The swabs for VRD were immediately stored dry in sterile test tubes and stored and transported on dry ice until processing at the laboratory. The VRD were performed by trained laboratory technicians within 6 hours after obtainment of the sample. Samples were left 15 minutes at room temperature before analysis in accordance with the manufacturer’s operating procedure. Test results were read visually after 15 minutes and thereafter with the VRD Analyser (VA).

Information on the first day of illness was subtracted from the GGD files for source and contact tracing.

In the second part of the study, participants were visited at home by GGD employees within 72 hours after their initial positive qRT-PCR at the GGD test centre. Analogous to the procedure in part one of the study, specimens for both qRT-PCR and VRD were obtained, stored and analysed. In addition, participants were asked what the day of symptom onset was and whether they still had symptoms at the time of the home visit.

### Sample size

We aimed to include 300 qRT-PCR negative individuals for part one of the study and 100 qRT-PCR positive individuals for the second part of the study.

### Performance analysis

For part one of the study the primary outcome was the VRD clinical sensitivity and specificity compared to qRT-PCR. For the second part of the study the primary outcome was VRD clinical sensitivity compared to qRT-PCR stratified by Ct-value category (Ct<20, Ct20-25, Ct25-30 and Ct≥30) and time since symptom onset (< 7 days, ≥ 7 days). Furthermore, overall positive predictive value (PPV) and negative predictive value (NPV) were calculated for a range of population prevalence (0.1%, 1.0%, 5%, 10%). In both parts of the study concordance between VRD analysis with the VA and visual interpretation were evaluated and compared with qRT-PCR results.

All data were analysed using Excel and SPSS version 24.

## Results

In part one of the study 354 individuals who presented at the test centre were initially included. 2 (0,6%) specimens with a negative VRD result were excluded as the result of qRT-PCR could not be recovered because of an error in sample number registration. 17 samples had detectable SARS-CoV-2 RNA, resulting in a prevalence of 4.8 per 100 participants. Amongst the 17 qRT-PCR positive specimen 12 (70,6%) were obtained within 7 days after disease onset, one (5,9%) was obtained later and for four specimens (23,5%) the time since symptom onset could not be determined. One qRT-PCR negative specimen rendered an uninterpretable and invalid VRD result by respectively visual interpretation and interpretation with the analyser. VRD was positive for 16 specimens based on visual interpretation and for 18 specimens based on interpretation with the analyser. The two samples which were positive with the analyser and negative by visual reading had a negative qRT-PCR. All 16 samples which were positive based on visual interpretation were qRT-PCR positive. Specificity was 100% (95%CI: 98,9%-100%) based on visual interpretation and 99,4% (95%CI: 97,9%-100%) based on interpretation with the analyser, the sensitivity was 94,1% (95%CI: 71,1%-100%) (table 1). The single qRT-PCR positive sample that was tested negative with VRD had a Ct-value of 32,7 and unknown time since symptom onset.

**Table 1:** VRD performance compared to qRT-PCR in Study Part 1.

|  | Visual interpretation | Interpretation with analyser |
| --- | --- | --- |
| Total (n) | 352 | 352 |
| Invalid | 1 | 1 |
| True positive (n) | 16 | 16 |
| False positive(n) | 0 | 2 |
| True negative (n) | 334 | 332 |
| False negative (n) | 1 | 1 |
| Sensitivity (%) [95% CI] | 94,1% [71,1%-100%] | 94,1% [71,1%-100%] |
| Specificity (%) [95% CI] | 100% [98,9%-100%] | 99,4% [97,9%-100%] |

In part two of the study, 132 individuals were included of which 129 (97,7%) had been or were symptomatic at the time of the home visit. The three asymptomatic individuals were tested in context of contact tracing (high-risk contact with a proven qRT-PCR positive individual). One was tested qRT-PCR and VRD positive, one qRT-PCR positive and VRD negative and one was tested negative in both. Six (4,5%) symptomatic individuals had a negative qRT-PCR at time of the home visit, all had a negative VRD. Clinical sensitivity of the VRD in symptomatic individuals was 78,9% (95%CI: 70,6%-85,7%). All but one Ct-value of the positive patients were obtained by the Roche 6800 qRT-PCR. The one exception which was tested on the Abbott platform and had a Ct-value below 20. When stratified by Ct-value category, sensitivity was found to be the highest in the lower Ct-value categories (highest viral loads) (Ct<20 100% (95%CI: 83,2%-100%), Ct20-25 93,3% (95%CI: 81,7%-98,6%), Ct25-30 88,2% (95%CI: 72,6%-96,7%), Ct>30 20,8% (95%CI: 7,1%-42,2%)). When subdivided in time since symptom onset shorter than 7 days or 7 days or more, clinical sensitivity was the highest for those specimens obtained within 7 days after symptom onset overall and for every Ct-value category (table 2).

**Table 2:** Test results of 123 qRT-PCR positive specimen of symptomatic individuals from Study Part 2.

| Days since symptom onset | Ct-value category | qRT-PCR + samples (n) | VRD + (n) | VRD - (n) | Sensitivity (%) [95% CI] |
| --- | --- | --- | --- | --- | --- |
| < 7 days | Ct < 20 | 17 | 17 | 0 | 100% [80,1%-100%] |
|  | Ct 20-25 | 29 | 29 | 0 | 100% [88,1%-100%] |
|  | Ct 25-30 | 12 | 11 | 1 | 91,7% [61,5%-99,8%] |
|  | Ct ≥ 30 | 8 | 2 | 6 | 25,0% [3,2%-65,1%] |
|  | Overall | 66 | 59 | 7 | 89,4% [79,4%-95,6%] |
|  | **CT < 30** | **58** | **57** | **1** | **98,3% [90,8%-100%]** |
| ≥ 7 days | Ct < 20 | 3 | 3 | 0 | 100% [29,2%-100%] |
|  | Ct 20-25 | 16 | 13 | 3 | 81,3% [54,4%-96,0%] |
|  | Ct 25-30 | 22 | 19 | 3 | 86,4% [65,1%-97,1%] |
|  | Ct ≥ 30 | 16 | 3 | 13 | 18,8% [4,1%-45,7%] |
|  | Overall | 57 | 38 | 19 | 66,7% [52,9%-78,6%] |
|  | **CT < 30** | **41** | **35** | **6** | **85,4% [70,8%-94,4%]** |

Combined, study part one and two comprised 140 qRT-PCR positive samples of symptomatic individuals. The time since symptom onset could not be determined for 4 qRT-PCR positive specimens of part 1 of the study, all but one were tested positive with VRD. Overall clinical sensitivity was 80,7% (95% CI: 73,2%-86,9%) with a sensitivity of 91.0% (95% CI: 82,4%-96,3%) for specimen (n=78) obtained within 7 days after disease onset. Sensitivity was found to be the highest in samples with high SARS-CoV2 viral loads (based on Ct-value) and for samples obtained early after the time of symptom onset. (figure 1) Sensitivity within 7 days after symptom onset was 100% (95% CI: 93,7%-100%) for qRT-PCR positive samples with a Ct-value of 25 or smaller and 98,6% (95% CI: 92,3%-100%) for Ct-values beneath 30 (table 3).

**Table 3:** Test results of qRT-PCR positive specimen of symptomatic individuals for who the time since symptom onset was known from Study part 1 and 2 combined. (n=136)

| Days since symptom onset | Ct-value category | qRT-PCR + samples (n) | VRD + (n) | VRD - (n) | Sensitivity (%) [95% CI] |
| --- | --- | --- | --- | --- | --- |
| < 7 days | Ct < 20 | 23 | 23 | 0 | 100% [85,2%-100%] |
|  | Ct 20-25 | 34 | 34 | 0 | 100% [89,7%-100%] |
|  | Ct 25-30 | 13 | 12 | 1 | 92,3% [64,0%-99,8%] |
|  | Ct ≥ 30 | 8 | 2 | 6 | 25,0% [3,2%-65,1%] |
|  | Overall | 78 | 71 | 7 | 91,0% [82,4%-96,3%] |
|  | **CT < 30** | 70 | 69 | 1 | 98,6% [92,3%-100%] |
| ≥ 7 days | Ct < 20 | 3 | 3 | 0 | 100% [29,2%-100%] |
|  | Ct 20-25 | 16 | 13 | 3 | 81,3% [54,4%-96,0%] |
|  | Ct 25-30 | 23 | 20 | 3 | 87,0% [66,4%-97,2%] |
|  | Ct ≥ 30 | 16 | 3 | 13 | 18,8% [4,1%-45,7%] |
|  | Overall | 58 | 39 | 19 | 67,2% [53,7%-79,0%] |
|  | **CT < 30** | 42 | 36 | 6 | 85,7% [71,5%-94,6%] |

**Figure 1:**
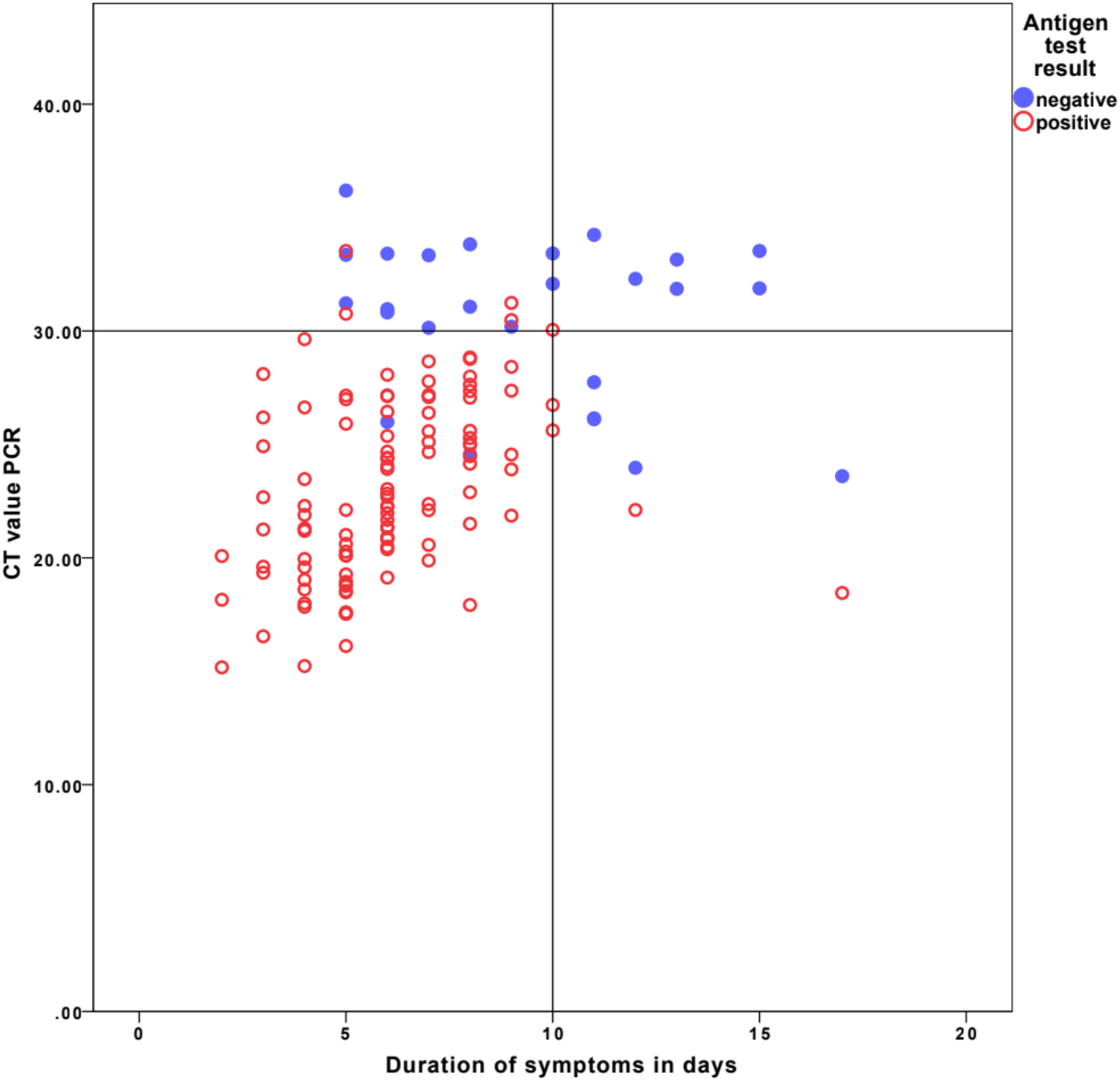
VRD results of 136 qRT-PCR positive specimen with known time of symptom onset (Part 1 and Part 2 combined) plotted against duration of symptoms (days) and qRT-PCR Ct-value.

Positive predictive value was always 100% as specificity was 100%. The negative predictive value for subjects who had symptoms since less than 7 days and a CT-value of 30 was above 99% up to a test-population prevalence of 60%. For the overall group who had symptoms less than 7 days, the negative predictive values for population prevalence’s of 10%, 20% and 50% were 99%, 98% and 95% respectively.

## Discussion

We found an overall clinical specificity of 100% (95% CI: 98,9%-100%) and sensitivity of 80,7% (95% CI: 73,2%-86,9%) of the VRD compared to qRT-PCR. Sensitivity was the highest for low Ct-value categories and for specimen obtained within the first days after disease onset. This is shown in figure 1. For specimen obtained within 7 days after onset of symptoms the sensitivity was 91,0% (95% CI: 82,4%-96,3%) overall and 98,6% (95% CI: 92,3%-100%) for samples with qRT-PCR Ct-value beneath 30.

To our knowledge no independent validation reports on the VRD have been published to date. Although numerous SARS-CoV-2 antigen tests developed by different manufacturers are available on the market, data on test performance is scarce. A recent review identified 5 performance evaluation studies evaluating a total of 8 SARS-CoV-2 antigen tests. The reported average specificity of 99.5% (95% CI: 98.1% to 99.9%) was in line with the results of our study. Sensitivity varied strongly across studies (from 0% to 94%) with an average of 56.2% (95% CI: 29.5% - 79.8%). The included studies were performed on remnant specimen stored in virus transport medium and often contained little information on days since disease onset and the clinical setting they were obtained in, all possibly explaining the discrepancy with the observed clinical sensitivity of the VRD in this study (5). Preliminary results of two performance evaluation studies of the Panbio Antigentest (Abbott) with to this study similar protocols performed on a total of 1397 samples were largely in line with the results observed in our study: an overall specificity of 100% and sensitivity of 73.2% (Utrecht) and 81.8% (Aruba). Similar to the VRD, the Panbio Antigentest was reported to perform better for lower Ct-value categories. (6)

The large sample size and the obtainment of samples in the target setting of potential use are great assets of the study. Furthermore, easy to perform superficial nose/throat swabs were used instead of difficult to perform and often performer dependent nasopharyngeal swabs.

As waiting times to make an appointment at a COVID-19 test centre were long during the study period due to the great demand, no specimen collected within 2 days after disease onset could be included. Although we expect this group to have high viral loads we cannot ascertain this assumption. The lack of data on this early window is a limitation of the study. COVID-19 infectivity peaks during the period shortly before and after the onset of symptoms when also maximal viral loads in upper respiratory tract material are measured. (7,8) In this context the test performance for specimen with a qRT-PCR Ct-value beneath 30 was calculated. As this cut off was based on the obtained data, it is to be confirmed by prospective evaluations.

In order to optimise standardisation, specimens were transported to the laboratory where the VRD was performed by trained technicians. As the final objective is to perform the VRD at the COVID-19 test centres, further research on test accuracy in point of care setting is needed. In order to perform the VRD at the laboratory, samples needed to be stored and transported on dry ice in accordance with the manufacturers’ prescriptions. Partial destruction of antigen due to freezing cannot be excluded and could have resulted in an underestimation of the clinical test sensitivity.

The presence of COVID-19 like symptoms is a pre-requisite to be tested at a GGD test-centre. As clients make their own appointment trough a digital system, we cannot exclude a small number of asymptomatic individuals amongst the included individuals in part one of the study. In part two of the study three asymptomatic subjects were excluded.

The current gold standard for diagnosis of an active SARS-CoV-2 infection is qRT-PCR. This highly sensitive and specific test is optimal for the diagnosis of clinically ill patients with a possible indication for treatment and individuals working in our staying at high-risk settings for outbreaks with severe consequences (e.g. long-term care facilities and hospitals). qRT-PCR is however expensive, has a long turnaround time (TAT) and can only be performed in specialised laboratories by trained personnel. The use of qRT-PCR for screening of community-dwelling individuals for the purpose of disease control creates an immense demand, leading to pressure on microbiologic laboratories compromising their routine clinical activities. Furthermore, logistic and administrative challenges intrinsic to qRT-PCR characteristics lead to substantial waiting times to get tested and to receive results. Rapid testing and feedback are however essential for control of SARS-CoV-2 community spread. (2) SARS-CoV-2 community screening for the purpose of disease control requires a low-cost, rapid diagnostic test that can be performed close to the community. Lateral flow SARS-CoV-2 antigen tests (LAF) can be performed at point of care, give results after 15-30 minutes and are less complex and expensive to produce. (3,4)

For subjects with complaints that started less than 7 days before testing, the negative predictive value was 98% for a test-population with a 20% prevalence. This value increases when the test-population prevalence becomes lower. At the time of writing, a second wave of COVID-19 infections was seen in the Netherlands with a prevalence of 10% to 20% in the test populations. In a questionnaire performed by the Dutch National Institute for Public Health and the Environment (RIVM) amongst 50.000 citizens in June 2020 only 12% of the interviewees that developed symptoms reported to have been tested. (9) The United States Centers for Disease Control and Prevention (CDC) estimated in June 2020 that only 10% of the COVID-19 cases in the United States were detected. (10) When 10% of COVID-19 infected individuals are tested with a 100% sensitive test, 900 in 1000 infected individuals will remain undetected. This strongly supports the use of additional tests with slightly lower sensitivity. We believe the beneficial effect of optimising test accessibility, as well geographically as in time, on the willingness to get tested will outweigh the limited decrease in test sensitivity by far.

Furthermore, COVID-19 infectivity and viral load in the upper respiratory tract generally peak around the time of symptom onset and decrease gradually during the following days. (8,11) Infected individuals should be detected in this first timeframe in order to optimise the effect of quarantine measures and contact tracing. For the purpose of COVID-19 control, it is preferential to test early on with a test with suboptimal analytical sensitivity for low viral loads, rather than using a 100% sensitive test only later on in the disease process.

In conclusion, the VRD is a promising diagnostic test for COVID-19 community screening for symptomatic individuals within 7 days after symptom onset in function of disease control. Performance of the test on a large scale is however likely to impose specific logistic challenges. Furthermore, the optimal position of the test within the current testing landscape is to be determined. Among others, further research is required to study the tests’ practical applicability and to determine the populations, test indications and settings in which it is most suited.

## Data Availability

Anonimised crude data is present at the Microvida laboratory for Microbiology in Breda, The Netherlands and will be saved.

## Transparency Declaration and Acknowledgements

Becton, Dickinson and Company was not actively involved in the design, execution, analysis or result interpretation of the study.

The VRD tests for this study were provided by the Dutch Ministry of Health, Welfare and Sport (VWS). We want to thank the employees of the Microvida laboratory for Medical Microbiology at the Amphia Hospital Breda and the GGD West-Brabant for their substantial contribution to this study.

Jan Kluytmans is member of the National Outbreak Management Team of The Netherlands and of a committee which supports the implementation of the Corona-reporting App.

